# Functional Disability and Predictors of Post-Intensive Care Syndrome During the Subacute Recovery Phase In a Middle-Income Country

**DOI:** 10.1101/2025.11.06.25339739

**Authors:** Chutima Cheranakhorn, Nichakan Nakwan, Suratee Chobngam, Sorawat Sangkeaw, Thitikorn Jongjaroenwit, Chananya Changadwej

## Abstract

**Background:** Post-Intensive Care Syndrome (PICS) contributes substantially to long-term disability among ICU survivors, although evidence from low- and middle-income countries (LMICs) remains limited. This study examined the prevalence and predictors of physical PICS during the subacute recovery phase in Thailand, representing a middle-income healthcare context.

**Methods:** A cross-sectional study was conducted among adult medical-ICU survivors at a tertiary hospital between March 2024 and February 2025. Patients admitted > 48 hours and surviving to hospital discharge were assessed for functional status at 3–9 months post-discharge using the Simplified Barthel Index (SBI). Univariable and multivariable logistic regression analyses identified independent predictors of physical PICS.

**Results:** Of 94 participants, 21 (21.3 %) met criteria for physical PICS. The median age was 59.5 years, and patients with PICS were significantly older than those without (72 vs 56 years, p < 0.01). Independent predictors included older age (aOR 1.07 per year, 95 % CI 1.02–1.13), prolonged mechanical ventilation (aOR 2.07 per day, 95 % CI 1.21–3.56), and longer hospital stay (aOR 1.17 per day, 95 % CI 1.02–1.34). Female sex was associated with higher risk (aOR 0.18; 95 % CI 0.03–0.99), whereas hyperglycemia showed an inverse relationship (aOR 0.08; 95 % CI 0.01–0.57).

**Conclusions:** Physical PICS affected one-fifth of ICU survivors during the subacute phase and was associated with advanced age, prolonged ventilation, and extended hospitalization. These findings underscore the need for early rehabilitation strategies and long-term follow-up systems for ICU survivors, particularly in LMIC settings where survivorship resources remain limited.

**Key Messages:**

- Post-Intensive Care Syndrome (PICS) contributes to long-term physical disability among ICU survivors, but data from low- and middle-income countries (LMICs) are limited.
- This study examined physical PICS during the subacute recovery phase (3–9 months post-discharge)—a period rarely explored in global literature.
- Older age, prolonged mechanical ventilation, and extended hospital stay were identified as independent predictors of post-ICU disability.
- Male sex appeared protective, while hyperglycemia reflected metabolic patterns specific to the study population rather than a causal factor.
- These findings emphasize the need for structured post-ICU follow-up and rehabilitation systems to improve survivorship outcomes in resource-limited healthcare settings.

## Introduction

Post-Intensive Care Syndrome (PICS) encompasses new or worsening physical, cognitive, and psychological impairments that persist beyond critical illness, affecting survivors’ independence, quality of life, and long-term mortality^1,2^. Reported prevalence varies widely: physical disability affects 25–80% of ICU survivors, cognitive impairment 30–80%, and mental health disorders 8–57%^3^. These sequelae collectively represent a growing global health burden, particularly as ICU survival rates improve.

While evidence from high-income countries has established PICS as a major public health challenge, evidence from low- and middle-income countries (LMICs) remains limited, despite the rapid expansion of critical care capacity. In Thailand, a study from a university hospital reported a prevalence of 50% in the physical domain, 27% in cognition, and 16% in mental health within 1–3 months after ICU discharge^4^. A national cohort of 7,070 ICU survivors demonstrated 5-year mortality of 35.7%, more than double that of age- and sex-matched controls^5^. Internationally, a multicenter study from Japan further demonstrated that the presence of PICS was strongly associated with excess mortality (PICS vs. non-PICS, 54% vs. 94% survival, p<0.01)^6^.

Despite these findings, most critical care research continues to emphasize short-term survival rather than long-term functional recovery. Systematic follow-up and rehabilitation services are rarely integrated into ICU care pathways, particularly in LMICs where resource limitations and fragmented post-discharge systems exacerbate the burden of disability. Furthermore, most existing studies have focused on the early recovery phase (1–3 months), whereas the subacute phase (3–9 months), a critical period for either regaining independence or progressing to chronic disability, remains underexplored.

This study aimed to examine the prevalence and predictors of physical PICS during the subacute recovery phase among medical ICU survivors in Thailand, providing new evidence from a middle-income healthcare context to inform post-ICU survivorship strategies globally.

## Methods

### Study Design and Setting

This cross-sectional study was conducted in the 43-bed medical intensive care unit (ICU) of Hatyai Hospital, a 1,033-bed tertiary-care referral center in southern Thailand. Adult ICU survivors admitted between March 1, 2024, and March 1, 2025, were included.

### Participants

Eligible participants were patients aged ≥18 years who had been admitted to the medical ICU for more than 48 hours and survived to hospital discharge. Exclusion criteria included post-cardiac arrest status, pregnancy, pre-existing severe disability (bedridden prior to admission), and palliative or end-of-life care.

### Data Collection

Demographic and clinical data were obtained from electronic medical records, including age, sex, comorbidities, admission diagnosis, Acute Physiology and Chronic Health Evaluation II (APACHE II) score, laboratory parameters, duration of intubation, ICU and hospital lengths of stay, and in-hospital complications such as hospital-acquired infections. Physical functional status was assessed using the Simplified Barthel Index (SBI)¹³ at 3–9 months after discharge.

### Outcome Definition

The primary outcome was physical PICS, defined as an SBI score ≤18¹⁴ at 3–9 months post-discharge, representing moderate to severe dependence in activities of daily living.

### Sample Size Calculation

The sample size was estimated using the single population proportion formula: 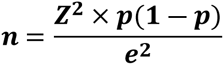 where *Z* represents the standard normal value corresponding to the desired confidence level, *p* is the expected prevalence, and *e* is the acceptable margin of error. Using an expected prevalence of 50% (based on a previous Thai study^4^), a 90% confidence level (*Z* = 1.645), and a margin of error of 9.2% (*e* = 0.092*), the calculated sample size was 84. Allowing for a 10% anticipated dropout, the final sample size was 93 participants, which matched the number enrolled in this study.

### Statistical Analysis

All analyses were performed using R software, version 4.0 (R Foundation for Statistical Computing, Vienna, Austria). Categorical variables were summarized as frequencies and percentages, and continuous variables as mean ± standard deviation (SD) or median (interquartile range, IQR), depending on data distribution. Group comparisons were conducted using the Chi-square or Fisher’s exact test for categorical data and the independent t-test or Mann–Whitney U test for continuous data. Variables with *p* < 0.05 in univariable analysis were entered into a multivariable logistic regression model to identify independent predictors of physical PICS. Results were expressed as adjusted odds ratios (aOR) with 95% confidence intervals (CIs), and statistical significance was set at *p* < 0.05.

### Ethics Statement

The study protocol was reviewed and approved by the Ethics Committee of Hatyai Hospital, Songkhla, Thailand (EC No. [HYH EC 002-66-01]). All procedures were conducted in accordance with the Declaration of Helsinki. Informed consent was waived because the study was retrospective and used anonymized data.

## Results

During the 12-month study period (March 1, 2024–March 1, 2025), 541 patients were admitted to the medical ICU. After exclusions, 94 survivors were included (Figure 1). Among them, 20 patients (21.3%) met criteria for physical PICS. The median age was 59.5 (39.2–70.8) years, and patients with PICS were significantly older [72 (65.5–83.2) vs. 56 (39–67.5) years, *p* = 0.002]. Males accounted for 58.5% of all participants, with no significant sex difference between groups. The mean APACHE II score was 17.5 ± 6.7. The predominant admission diagnoses were infection other than pneumonia (34%) and pneumonia (10.6%), tuberculosis was more common in the PICS group (10% vs. 0%, *p* = 0.043). The leading comorbidities were diabetes (37.2%), hypertension (36.2%), and dyslipidemia (25.5%), without intergroup differences.

**Figure 1:**
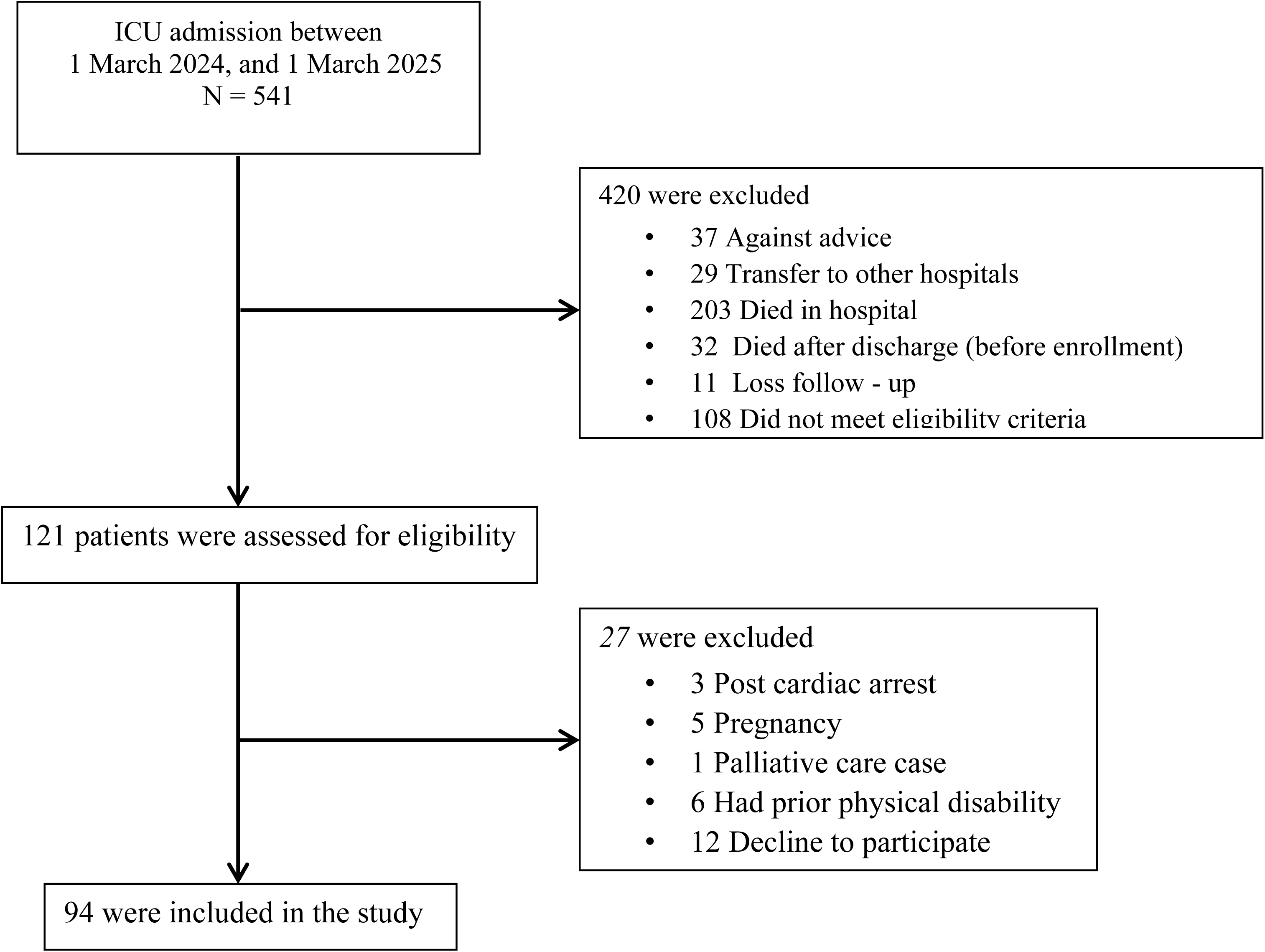
Patient flow diagram. Flow of patient enrollment, exclusion, and inclusion between March 2024 and March 2025.

A total of 83% of patients required mechanical ventilation; the median intubation duration was significantly longer in the PICS group [8.5 (3–12) vs. 2 (1–4) days, *p* < 0.001]. Hospital-acquired infection occurred more frequently among PICS patients (60% vs. 10.8%, *p* < 0.001). The median hospital stay was longer in the PICS group [19.5 (13–31.5) vs. 7.5 (5.2–12) days, *p* < 0.001], as was the ICU stay [9.5 (4–13.2) **vs.** 3 (2–5) days, *p* < 0.01] (Table 1)

**Table 1:** Patient characteristics.

| Characteristic | All (N=94) | PICS group (N=20) | Non-PICS group (N=74) | *p*-value |
| --- | --- | --- | --- | --- |
| Age – yr, median (IQR) | 59.5 (39.2-70.8) | 72 (65.5-83.2) | 56 (39-67.5) | 0.002 |
| Male sex – no. (%) | 55 (58.5) | 8 (40) | 47 (63.5) | 0.058 |
| Diagnosis – no. (%) |  |  |  |  |
| Pneumonia | 10 (10.6) | 3 (15) | 7 (9.5) | 0.439 |
| Other infections | 32 (34) | 4 (20) | 28 (37.8) | 0.135 |
| Upper GI bleeding | 10 (10.6) | 1 (5) | 9 (12.2) | 0.683 |
| DKA/HSS | 9 (9.6) | 1 (5) | 8 (10.8) | 0.679 |
| Heart failure | 9 (9.6) | 2 (10) | 7 (9.5) | 1.000 |
| COPD/Asthma | 5 (5.3) | 2 (10) | 3 (4.1) | 0.287 |
| Acute ischemic stroke | 1 (1.1) | 1 (5) | 0 (0) | 0.213 |
| Tuberculosis | 2 (2.1) | 2 (10) | 0 (0) | 0.043 |
| Other | 15 (16) | 4 (20) | 11 (14.9) | 0.731 |
| Underlying disease – no. (%) |  |  |  |  |
| Diabetes mellitus | 35 (37.2) | 7 (35) | 28 (37.8) | 0.816 |
| Hypertension | 34 (36.2) | 7 (35) | 27 (36.5) | 0.902 |
| Dyslipidemia | 24 (25.5) | 2 (10) | 22 (29.7) | 0.073 |
| Chronic kidney disease | 12 (12.8) | 3 (15) | 9 (12.2) | 0.714 |
| Heart disease | 10 (10.6) | 4 (20) | 6 (8.1) | 0.212 |
| COPD/Asthma | 9 (9.6) | 3 (15) | 6 (8.1) | 0.395 |
| Cirrhosis | 6 (6.4) | 0 (0) | 6 (8.1) | 0.336 |
| Malignancy | 6 (6.4) | 1 (5) | 5 (6.8) | 1 |
| Cerebrovascular disease | 3 (3.2) | 2 (10) | 1 (1.4) | 0.113 |
| Other neurological disease | 5 (5.3) | 2 (10) | 10 (13.5) | 0.287 |
| HIV infection | 3 (3.2) | 1 (5) | 2 (2.7) | 0.516 |
| Other | 12 (12.8) | 2 (10) | 10 (13.5) | 1.000 |
| Initial laboratory testing |  |  |  |  |
| Hematocrit – mean (SD) | 31.4 (9.1) | 31.9 (7.2) | 31.3 (9.6) | 0.804 |
| Platelet – median (IQR) | 219,000<br>(105,250-293,500) | 259,000<br>(8,947-14,885) | 204,500<br>(97,000-285,750) | 0.017 |
| Creatinine – median (IQR) | 1.2 (0.8-2.4) | 1.1 (0.6-1.9) | 1.2 (0.8-2.5) | 0.237 |
| pH – median (IQR) | 7.4 (7.3-7.4) | 7.4 (7.3-7.4) | 7.4 (7.3-7.4) | 0.674 |
| PaCO<sub>2</sub> – median (IQR) | 25.6 (20-31.8) | 28.3 (23.7-40.5) | 25 (19.2-31.4) | 0.11 |
| Lactate – median (IQR) | 2.3 (1.1-4.5) | 2.2 (1.4-3.9) | 2.3 (1.1-4.6) | 0.923 |
| Albumin – mean (SD) | 3.3 (0.7) | 3.2 (0.8) | 3.3 (0.7) | 0.902 |
| APACHE II score – mean (SD) | 17.5 (6.7) | 18.1 (5.2) | 17.3 (7.1) | 0.682 |
| Intubation – no. (%) | 78 (83) | 18 (90) | 60 (81.1) | 0.508 |
| Intubation days – median (IQR) | 2 (1-5) | 8.5 (3-12) | 2 (1-4) | <0.001 |

**Table 1: Patient characteristics (continue)**
| Characteristic | All (N=94) | PICS group (N=20) | Non-PICS group (N=74) | *p*-value |
| --- | --- | --- | --- | --- |
| Criteria for ICU admission – no. (%) |  |  |  |  |
| Shock | 43 (45.7) | 8 (40) | 35 (47.3) | 0.561 |
| Respiratory support | 83 (88.3) | 19 (95) | 64 (86.5) | 0.447 |
| Renal replacement | 10 (10.6) | 2 (10) | 8 (10.8) | 1 |
| Post operative care | 2 (2.1) | 0 (0) | 2 (2.7) | 1 |
| Hemodynamic monitor | 6 (6.4) | 2 (10) | 4 (5.4) | 0.604 |
| Special procedure | 1 (1.1) | 1 (5) | 0 (0) | 0.213 |
| Shock – no. (%) |  |  |  | 0.713 |
| Septic shock | 30 (71.4) | 7 (87.5) | 23 (67.6) |  |
| Hemorrhagic shock | 5 (11.9) | 0 (0) | 5 (14.7) |  |
| Cardiogenic shock | 2 (4.8) | 1 (12.5) | 1 (2.9) |  |
| Adrenal crisis | 2 (4.8) | 0 (0) | 2 (5.9) |  |
| Obstructive shock | 1 (2.4) | 0 (0) | 1 (2.9) |  |
| Hypovolemic shock | 1 (2.4) | 0 (0) | 1 (2.9) |  |
| Cardiac tamponade | 1 (2.4) | 0 (0) | 1 (2.9) |  |
| Hospital acquired infection – no. (%) | 20 (21.3) | 12 (60) | 8 (10.8) | < 0.01 |
| Renal replacement therapy – no. (%) | 15 (16) | 2 (10) | 13 (17.6) | 0.513 |
| Volume overload | 7 (46.7) | 1(50) | 6 (46.2) |  |
| Metabolic acidosis | 5 (33.3) | 1 (50) | 4 (30.8) |  |
| Uremia | 2 (13.3) | 0 (0) | 2 (15.4) |  |
| Anuria | 1 (6.7) | 0 (0) | 1 (7.7) |  |
| Dysglycemia during admit– no. (%) |  |  |  |  |
| Hypoglycemia | 5 (5.3) | 2 (10) | 3 (4.1) | 0.287 |
| Hyperglycemia | 43 (45.7) | 7 (35) | 36 (46.8) | 0.277 |
| Delirium – no. (%) | 9 (9.6) | 2 (10) | 7 (9.5) | 1 |
| Corticosteroid use – no. (%) | 27 (28.7) | 5 (25) | 22 (29.7) | 0.678 |
| Neuromuscular blocking agent use – no. (%) | 3 (3.2) | 0 (0) | 3 (4.1) | 1 |
| Sedative drug use – no. (%) | 11 (11.7) | 2 (10) | 9 (12.2) | 1 |
| Length of hospital stay – median (IQR) | 9 (6-14.8) | 19.5 (13-31.5) | 7.5 (5.2-12) | <0.001 |
| Length of ICU stay – median (IQR) | 4 (2-7) | 9.5 (4-13.2) | 3 (2-5) | <0.01 |

IQR=interquartile range; COPD= chronic obstructive pulmonary disease; DKA= diabetic ketoacidosis; HHS=Hyperosmolar hyperglycemic state; HIV= Human immunodeficiency virus; SD=standard deviation; APACHE Acute Physiology and Chronic Health Evaluation, ICU= intensive care unit

In the multivariable logistic regression model, three factors remained independently associated with physical PICS: older age (aOR 1.07; 95% CI 1.02–1.13; *p* = 0.01), longer duration of mechanical ventilation (aOR 2.07; 95% CI 1.21–3.56; *p* = 0.008), and longer hospital stay (aOR 1.17; 95% CI 1.02–1.34; *p* = 0.021). In addition, male sex was associated with a lower likelihood of developing physical PICS (aOR 0.18; 95% CI 0.03–0.99; *p* = 0.048), suggesting a potential protective effect compared with females. Hyperglycemia during ICU stay showed an inverse association (aOR 0.08; 95% CI 0.01–0.57; *p* = 0.012) (Table 2).

**Table 2:**
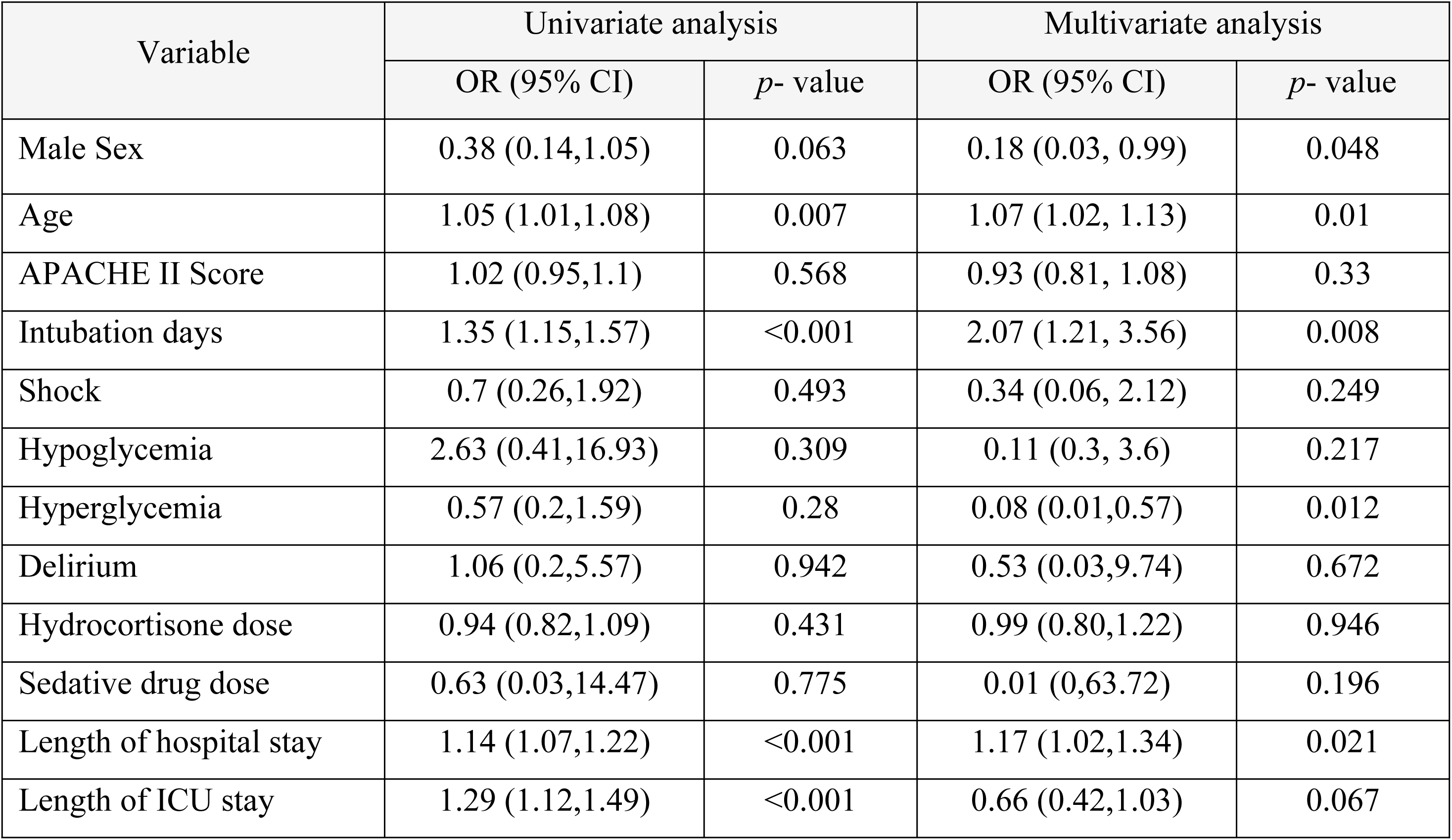
Univariate and Multivariate Logistic Regression Model of Factors Associated with Physical Disability Post-Intensive Care Syndrome.

## Discussion

In this study, the prevalence of physical post-intensive care syndrome (PICS) was 21%, lower than that reported in most previous studies. A recent systematic review and meta-analysis found a pooled global prevalence of 45.99% (95% CI 34.65–57.31) in the physical domain among ICU survivors^9^. Several factors may explain this discrepancy. First, the timing of follow-up likely influenced the observed prevalence. Earlier studies typical assessed patients within 1–3 months after ICU discharge, when disability tends to peak, whereas this study’s assessment at 3–9 months may have captured patients who had already regained partial function through rehabilitation or spontaneous recovery. Second, survivor bias may have reduced the measured prevalence, since patients with the most severe functional decline may have died before follow-up and were thus not represented. Third, the severity of illness appeared to correlate with PICS prevalence across studies. The present cohort (APACHE II = 17.5; estimated mortality 25–30%) had a physical PICS rate of 21%, indicating moderate illness severity and partial recovery. Similar findings were observed in Spain (APACHE II = 16.4; 32% PICS)^10^ and South Korea (APACHE II = 11.5; 19.8% PICS)^11^. In contrast, higher-acuity cohorts from Japan (APACHE II ≈ 20; 37% physical PICS at 6 months)^6^ and the Netherlands (APACHE IV = 62; ≈ APACHE II 17–20; mortality 20–30%) reported 57% physical PICS prevalence at 1 year^12^, suggesting that greater illness severity is associated with more persistent impairment and delayed recovery. Fourth, differences in measurement tools may contribute. The Simplified Barthel Index (SBI) was chosen for its simplicity, objectivity, and feasibility for telephone-based assessment, as well as its routine use at ICU and hospital admission and discharge, allowing direct comparison of functional status. Although the 6-Minute Walk Test is recommended by the Society of Critical Care Medicine, its low recommendation grade^13^ and limited feasibility in resource-constrained or non-ambulatory settings make the SBI a more pragmatic tool for large-scale or longitudinal follow-up studies. Finally, sociocultural factors may influence outcomes. In Thailand—particularly in rural and agricultural communities—extended families often live together across generations and provide mutual support in daily activities. This strong family and community support system may facilitate earlier recovery and adaptation, even without formal rehabilitation programs.

The downward trend in prevalence observed in this cohort aligns with the previously reported natural recovery trajectory, where PICS prevalence decreased from 55.4% at 3 months to 27.7% at 6 months^14^. Similarly, studies from South Korea, Japan, Spain, and the Netherlands reported that approximately half of ICU survivors experience some domain of PICS during the first year, with physical impairment ranging from 20– 50% and improving overtime^6, 10–12^. A prior Thai study from a university hospital further supports this trajectory, reporting a PICS incidence of 64.7% (50% physical, 27% cognitive, 16% psychological) within 28 days after discharge, with a mean APACHE II score of 16 ± 4.6^4^. The markedly lower prevalence observed in this cohort at 3–9 months suggests substantial recovery during the subacute phase of survivorship.

The multivariable analysis identified several independent predictors of physical PICS that were consistent with international data while providing regional insight from a middle-income healthcare context. Older age remained a strong predictor (aOR 1.07 per year, 95% CI 1.02–1.13), reflecting the recognized vulnerability of elderly patients to post-critical illness sequelae through reduced physiological reserve, pre-existing sarcopenia, and impaired recovery mechanisms^15–17^. Prolonged mechanical ventilation demonstrated the strongest association with physical PICS (OR 2.07 per day, 95% CI 1.21-3.56), consistent with evidence showing that diaphragmatic dysfunction begins within hours of mechanical ventilation, with measurable structural changes occurring within 12-24 hours and progressive muscle fiber atrophy throughout the duration of ventilatory support^18–20^. An extended hospital stay independently predicted physical disability (aOR 1.17 per day, 95% CI 1.02–1.34), likely representing cumulative exposure to deconditioning, nosocomial complications, and prolonged immobilization inherent to critical care recovery^21,22^. Male sex was associated with a lower risk of physical PICS, consistent with prior evidence showing that female survivors tend to have slower functional recovery and greater fatigue after critical illness^23^. This difference may reflect variations in baseline muscle mass, hormonal influences on muscle repair, and sociocultural factors affecting post-discharge rehabilitation^24^. Hyperglycemia showed an inverse association (aOR 0.08; 95% CI 0.01–0.57), likely reflecting the inclusion of younger diabetic ketoacidosis patients with reversible metabolic derangements and favorable recovery potential rather than stress hyperglycemia secondary to sepsis or systemic inflammation^25^.

Several limitations warrant consideration. This single-center observational study had a modest sample size, which may limit generalizability. The analysis focused on the physical domain during the subacute (3–9 months) phase; long-term outcomes and other PICS domains require further study. Nonetheless, this investigation offers new insight into post-ICU recovery in a real-world, resource-limited setting, where structured rehabilitation and survivorship programs remain scarce. By characterizing recovery trajectories within this context, the study contributes to addressing a global evidence gap and reinforces that established predictors, such as age, ventilation duration, and hospital stay, are robust across diverse healthcare systems. These findings emphasize the need to adapt and implement evidence-based practices, such as early mobilization, infection prevention, and multidisciplinary follow-up, to improve survivorship outcomes in all critical care environments, particularly in low- and middle-income countries.

## Conclusion

Physical post-intensive care syndrome (PICS) affected one in five ICU survivors in this cohort, with recovery patterns consistent with international data but assessed during the underexplored subacute recovery phase (3–9 months). Older age, prolonged mechanical ventilation, and extended hospitalization were key predictors of persistent functional impairment, whereas male sex appeared protective, and hyperglycemia reflected cohort-specific metabolic characteristics rather than a causal association. These findings highlight the importance of early recognition of high-risk patients and coordinated, multidisciplinary follow-up to sustain recovery after critical illness. The study also contributes to narrowing the global evidence gap on post-ICU trajectories and supports the incorporation of structured survivorship programs into contemporary critical care practice.

## Declarations

## Acknowledgments

The authors thank the medical and nursing staff of the Medical Intensive Care Unit, Hatyai Hospital, for their collaboration and continuous support throughout the study.

## Author Contributions

Chutima Cheranakhorn conceived the study, designed the protocol, and supervised data collection. Nichakan Nakwan and Suratee Chobngam contributed to data verification and methodological consultation. Sorawat Sangkaew performed statistical analyses and interpretation. Thitikorn Jongjaroenwit and Chananya Changadwej assisted with data collection and clinical follow-up. All authors participated in manuscript drafting and revision and approved the final version for publication.

## Data Availability

The datasets generated and analyzed during the current study are available from the corresponding author upon reasonable request.

